# Assessing adverse childhood experiences and mental health status in diverse and underrepresented young people: advancing Inclusive research

**DOI:** 10.64898/2026.07.21.26358568

**Authors:** Kamaldeep Bhui, Megan Kirk, Isabelle Butcher, Mina Fazel, Minhua Ma, Paul Cooke, Chloe Farahar, Annette Foster, Kier Harris, Harsimran Sansoy, Laura Havers, Nicola Shaughnessy, Siobhan Hugh-Jones, Luke Allder, Anna Mankee-Williams

## Abstract

**Background:** Young people impacted by adverse childhood experiences (ACEs) are often underrepresented in mental health research.

**Aims:** This paper aims to advance inclusive research on ACEs by 1) describing co-designed recruitment and engagement methods in a national project on ACEs (Attune), 2) characterising a highly marginalised cohort of young people using identity descriptors co-designed with participants, and 3) reporting associations between ACEs, identity characteristics and mental health outcomes.

**Methods:** A trauma-aware approach to engage under-represented young people was co-developed with a national youth advisory group, lived experience researchers, and trusted community partners. Our co-created purposive sampling strategy recruited 74 young people, aged 10-24 years, across England, seeking representation by age, sex, gender identity, sexual orientation, ethnicity, neurodivergence, and geographic location. Participants completed validated self-report measures of ACEs, life events, and mental health. Descriptive, correlational and regression analyses examined cohort characteristics and associations between ACEs, identity characteristics, and mental health measures.

**Results:** The final cohort included participants identifying as non-White British (39.5%), non-binary/other gender (25%), and neurodivergent (30%). Half of participants reported exposure to at least one ACE. Analyses identified patterns consistent with prior literature. In addition, ACEs and barriers related to being neurodivergent were associated with increased depression and anxiety symptom severity. Non-binary gender identity was associated with anxiety. We did not observe associations of ACEs or mental health measures, with sex or ethnicity.

**Conclusions:** Under-represented groups can be reached via co-created engagement methods informed by lived experience. We identified important associations between ACEs, identities, and mental health outcomes.

## Introduction

### Background

Mental health difficulties in young people increased during and following the COVID- 19 pandemic leading to increasing demands on services.^1^ These difficulties disproportionately affect the most marginalised by social identity or social position (e.g., by ethnicity, sexual and gender identity, socioeconomic circumstances, and neurodivergence) and those who are exposed to adverse childhood experiences (ACEs).^2^ ^3^ Abuse, neglect, and discrimination are well-established risk factors for poorer mental health across the life course.^4^ Because marginalised young people are also at heightened risk of exposure to ACEs,^5^ research into ACEs and associated mental health consequences must include and represent young people facing multiple disadvantages. Paradoxically, the most marginalised young people with the greatest needs are least well served by existing care systems (e.g., neurodivergent young people) and are typically under-represented in research seeking to improve care.^6^ ^7^ Recently, many researchers have attempted to understand what under-representation means for young people directly and the impact of biases in evidence that informs policy and practice.^8^ ^9^

Although representation is improving for specific characteristics such as younger age groups, by sex and by ethnicity, under-representation of complex groups with multiple intersecting vulnerabilities remains a challenge. Although this problem is recognised, more research is needed on how to improve marginalised young people’s involvement and retention in studies, especially in studies on ACEs which typically ask about sensitive and personal experiences. These studies can raise concerns about triggering and are often associated with understandable emotional and behavioural avoidance.^10^ Hence, the reasons for under-representation may include a lack of trust and worries about re-traumatisation, whereby ensuring safeguarding protections are in place, and working through community or parents can be helpful.^11^ ^12^ Additional barriers include insufficiently resources to adequately support participation, complex consent processes that are difficult to navigate for vulnerable young people, literacy demands, geographical isolation requiring elaborate travel arrangements, and living in precarity which makes any additional demands feel overwhelming.^13^ ^14^ Despite growing recognition of these reasons, there remains little applied evidence on how inclusive and trauma-aware principles can ensure safety and support the recruitment and representation of marginalised young people, especially in research on the mental health impacts of ACEs.

### Study purpose

The current paper reports on participants recruited to the first phase of ATTUNE, a mixed-methods, UK Research and Innovation (UKRI) funded project, to understand young people’s experiences of ACEs. The national programme sought to address the needs of adolescents aged 10-24 years, whilst acknowledging the variation in developmental stages and different vulnerabilities across this age span. Given the challenges of including marginalised young people in research, the project sought to develop inclusive, co-designed, and trauma aware research practices.

The invitation to participate in the first phase of the ATTUNE project made explicit that intention to involve young people in participatory arts workshops, data from which were to undergo qualitative analysis. Although the methods and findings of these workshops are not reported here, briefly, these included visual arts (e.g. photography, mask-making, collage), spatial practices (e.g. den-building), making animations and short films, movement and dance, music, drama, and creative writing. These were selected to broaden accessibility beyond conventional verbal interviews for later stages of the project (attuneproiect.com). Individuals wishing to participate first completed an online questionnaire (detailed below) in part to ensure we were aware of support needs and could offer adequate support. The online questionnaire was finalised after piloting and feedback from young people and community partners. This required adjustments to language, building trust in the research methods, and proposing different ways of introducing the research that prioritised psychological safety.

This paper sets out the co-design and inclusive recruitment processes. We then describe cohort characteristics, mental health indicators, and exposure to ACEs and other life events. We then present associations between diversity characteristics, exposure to ACEs, and mental health outcomes to replicate and extend the evidence base to marginalised groups and improve the credibility and transferability of inclusive research practices.

## Methods

### Co-design and collaboration with young people

The recruitment methods were co-designed in collaboration with young people and community partners to support safe recruitment; youth input was gathered through local regional and national young people advisory groups (YPAGs).^13^ There were 58 young people in local and national advisory groups (Oxford (10 people), Leeds (7 people), Cornwall (23 people), Kent (8 people), and a national group (of 10 people, run by Centre for Mental Health)). Young people’s advisory groups (YPAGs) met quarterly and as necessary to discuss the project design and delivery. The national YPAG met more frequently around conferences and dissemination events. Agenda items for discussion were distributed early, and young people were prepared before meetings where they had additional communication or sensory needs.

In initial YPAG sessions across rural, urban and coastal regions, young people were informed of the study purpose and aims. To ensure that the YPAG members felt that they could voice their opinions, a series of guidelines on how to run the meetings was produced by YPAG members. The collaborative work with the YPAGs led to the co-design of a study poster (Annex A) and a video produced by young people to promote the study (First Oxford Film- Oxford YPAG.mp4). Materials were refined iteratively over several weeks, with revisions discussed and agreed upon across YPAG meetings organised by local and national YPAG groups (see structure of YPAG groups Annex B).

For each process and document, YPAG members were invited to shape participant facing materials, including study information sheets, consent and assent forms, recruitment flyers and procedures for approaching potential participants. YPAG members emphasised the importance of clear, plain language and transparency about what participation would involve, including how sensitive topics such as ACEs and mental health would be addressed.

YPAGs members emphasised the importance of recruitment through trusted partners rather than open advertisements, via established community and youth groups and schools with existing connections. A named person acted as a point of contact. Study information materials were revised to reduce academic or clinical terminology, explicitly acknowledge the potential emotional impact of questions that were to be asked in an online questionnaire as part of the recruitment procedures (see Table 1).

**Table 1:**
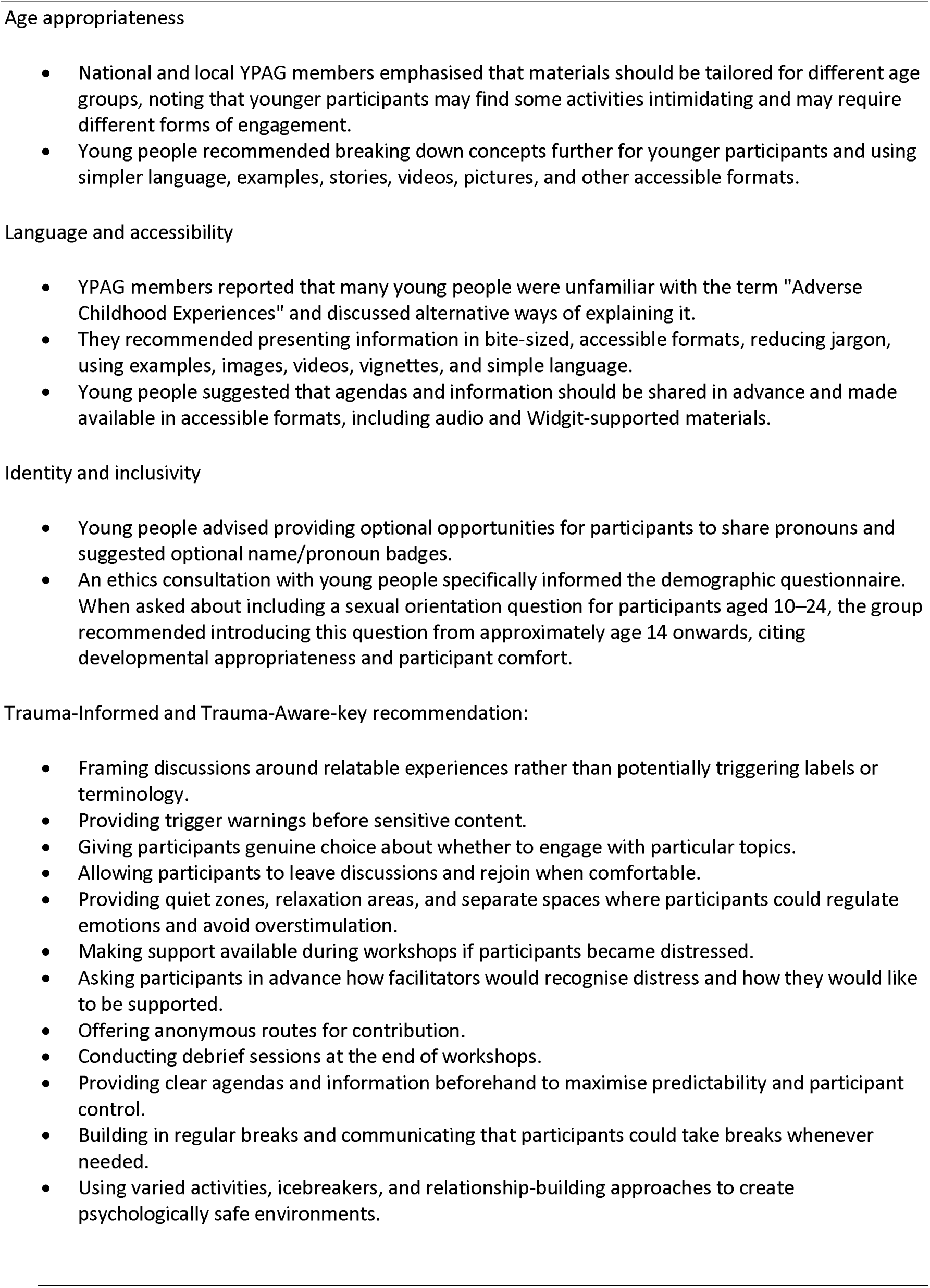
Adjustment advised by young people.

Young people highlighted concerns about standard demographic indicators that may not accurately reflect lived experience. For example, commonly used measures of socioeconomic status (e.g. number of cars in the household) were viewed as inappropriate in some rural contexts, where car ownership may be a necessity rather than a marker of affluence. YPAG members also revised the externally supplied wording of standard templates on diversity and response options used to describe gender identity, sexual orientation, ethnicity, and neurodivergence. This sought to avoid hierarchical, implicitly normative and invalidating categories. Care was taken to ensure that questions were inclusive, non-assumptive, and allowed participants to self-describe using free text where predefined categories felt insufficient. For neurodivergence, YPAGs advised distinguishing between self-identified (i.e., ‘I consider myself to be/have/experience’) and clinically diagnosed experiences, and allowing participants to select multiple neurodivergences.

Young people were consulted and invited to shape the content and delivery of demographic, ACE, and mental health questionnaires, as well as the questionnaires for additional outcomes of interest. We clearly stated participants’ rights to skip questions, pause, or withdraw at any point without explanation. YPAGs informed adaptations on sensitive wording (see Table 1). For example, the Short Childhood Maltreatment Questionnaire (SCMQ) asks participants to state whether they have experienced attempts by an adult to have ‘oral, anal or vaginal intercourse with [them]’. Consultation with two regional YPAGs reached consensus that this wording could feel explicit, confusing or retraumatising. The question was rephrased to: *Did someone at least five years older than you or an adult attempt or actually have unwanted intercourse with you?* This was to maintain conceptual equivalence while improving clarity and prevent risk of distress.

### Study participants

Recruitment focussed on five geographic regions of England: Cornwall, Kent, London, Oxfordshire, and Yorkshire, by purposive sampling of up to 15-20 young people from each region (total sample of 75-100) to ensure representation by age, sex, LGBTQ+, ethnicity, and neurodivergence.

### Study Procedures

Individuals were recruited through local community and charity organisations, as well as educational settings (listed in Annex C). Organisations were approached if they were known particularly to support marginalised young people, including those from the LGBTQ+ community, ethnically minoritised groups, Traveller communities, and neurodivergent individuals. In each organisation, a named lead person acted as the support, gatekeeping, and safeguarding official. We explained the purpose of the overall programme of work to all stakeholders, including young people who might wish to participate; however, we did not specifically recruit based on exposure to ACEs; rather we asked about these and other characteristics.

Ethics review of the entire research projects was concluded in 2022.The authors assert that all procedures contributing to this work comply with the ethical standards of the relevant national and institutional committees on human experimentation and with the Helsinki Declaration of 1975, as revised in 2013. All procedures involving human subjects/patients were approved by Oxford’s Research Ethics Committee (R71941/RE001) and the Health Research Authority (23/WM/0105). Assent was obtained for individuals below the age of 16, which included parental or carer informed written consent to participate. Informed written consent was obtained from individuals aged 16 and above.

A member of the research team was present throughout to provide support if needed, on completing consents. The lead from the partner organisation was also present. Privacy and data protection were assured through the online completion, and secure storage of data by personalised pseudonyms. Participants then completed a series of self-report questionnaires (see below) via the online platform or in person, and in the presence of a member of the ATTUNE research team, as suggested by YPAGs and partner organisations.

The questionnaire covered demographics, neurodivergences, ACEs, and mental health as well as additional relevant areas of wellbeing, loneliness, threatening life events, and emotion regulation. ^15^ ^16^ ^17^ ^18^ Participants were free to skip questions, ask for clarification, or leave the study without giving a reason. Post-study support, liaison with the partner organisations and with safeguarding leads was provided when required.

### Measures

#### Demographics

Participants completed a co-designed demographic questionnaire about their region of residence, age, sex, gender identity, sexual orientation, ethnicity, and religion. Items and response options aligned with the National Health Service (NHS) reporting standards in the UK, with additional options to select “prefer not to say” or to provide a free-text response. Participants were also asked about their experiences of care, their level of education, and current employment status. Socioeconomic circumstances were assessed using questions about their parent/caregiver/guardian’s education and occupation. Questions relating to sex, sexual orientation, and gender identity were co-developed with young people. YPAG facilitation included researchers with lived experience of neurodivergence and diverse gender/sexual identities, supporting relational trust and linguistic alignment during questionnaire development.

#### Neurodivergence

Participants were asked whether they self-identified as having one or more neurodivergences (including being Autistic, experiencing ADHD, and having a learning disability). Descriptive response options were co-designed with YPAGs and neurodivergent research team members. Participants were additionally asked whether the diagnosis was from a medical professional. There were multiple response options, and a free text box if their preferred terms were not. As outlined, YPAG members challenged medicalised and deficit-based language, leading to modifications wherever possible.

#### Adverse Childhood Experiences (ACEs)

Exposure to ACEs was assessed using the Short Child Maltreatment Questionnaire. The SCMQ was adapted from the World Health Organisation^19^ and includes seven items reflecting five dimensions of maltreatment: physical, emotional, sexual abuse, neglect, and witnessing parental physical violence. For each item participants are asked to indicate the presence and type of maltreatment experiences (“No, never”, “Yes, it happened in the last 12-months”, “Yes, it happened in my life”, “Prefer not to answer”), and the level of frequency and severity on a five-point response scale. Participants could report other adverse experiences. The original SCMQ includes two questions on acts of sexual violence. The wording for these items was adjusted following consultation with YPAGs to enhance acceptability (see results section). Total ACEs scores were derived and used in the statistical analyses.

#### Anxiety

Anxiety symptom severity was assessed using the Generalized Anxiety Disorder seven-item questionnaire (GAD-7)^20^. Participants rated how frequently they had experienced each symptom over the past two weeks. Responses were based on a 4-point scale ranging from ‘not at all’ to ‘nearly every day.’ Total scores (0-21) were used in the statistical analyses. Established cut-offs suggest that scores above 10 and 15 reflect moderate and severe anxiety, respectively.^21^

#### Depression

Depressive symptoms over the previous two weeks were measured using the Patient Health Questionnaire (PHQ-9).^21^ Items were rated on a four-point scale from “Not at all” to “Nearly every day” (Supplementary File 1). Total scores (0-27) were used in the statistical analyses, with cut-off scores above 10 and 20 interpreted as moderate and severe, respectively. Details of the secondary outcome measures used to assess the additional areas of threatening life events, wellbeing, loneliness, and emotion regulation, detailed in Annex C.^15^ ^18^

### Statistical analysis

Analyses were conducted using SPSS version 31.0.1. Data were cleaned, coded, and independently cross-checked by two investigators (MK, KB) prior to analysis. Demographic categories were recoded for analytic purposes where cell sizes were small. Gender was recoded from 6-item response options into 3 distinct categories (e.g., 1 = man/boy, 2 = woman/girl, 3 = non-binary/other (e.g., gender fluid). Sexual orientation was recoded from 9-item response options into 3 categories (i.e., 1 = LGBTQ+, 2 = straight/heterosexual, 3 = asexual/unsure). Ethnicity was recoded from 18-item response options into a dichotomous variable with 1 = White, and 2 = Non-White. Trans identity was recoded from 5-item response option into a dichotomous variable with 0 = No/Prefer not to say, and 1 = Yes/questioning/Unsure. Psychometric total scores for PHQ-9 and GAD-7 were used as the continuous dependent variable.

Initial descriptive analyses characterised the sample by ACEs, demographic and identity characteristics, and mental health outcomes. A bivariate correlation matrix was conducted to test initial associations between ACEs score, psychometric outcomes, and demographic variables to determine the direction and strength of associations between variables to help inform regression model testing. Pearson product-moment correlation coefficients were reported for continuous variable associations. For non-parametric data, Spearman rank-order and point-biserial correlations were conducted. A Pearson chi-square test and Phi or Cramer’s V contingency coefficients were calculated to assess the strength of association between two nominal variables. To test associations between demographic categories and identity characteristics with ACEs and mental health outcomes (e.g., PHQ-9, GAD-7), independent samples t-tests and one-way analyses of variance (ANOVAs) were conducted as appropriate, with Tukey’s HSD applied for post-hoc pairwise comparisons where applicable.

Initial descriptive analyses informed the regression model and variables entered for each primary outcome. Hierarchical multiple regression analyses were conducted to examine the independent associations between ACEs and selected demographic characteristics with either depression (PHQ-9) and anxiety (GAD-7) severity as the dependent variable. In Model 1, total ACEs score (i.e., SCMQ) was entered as the primary prediction variable. In Model 2, significant sociodemographic variables from initial analyses were entered to assess the additional variance explained in either PHQ-9 and GAD-7 scores. Gender identity (0 = man/boy, 1 = woman/girl, 2 = non-binary) was dummy coded using SPSS with dummy variables created for all values except the first category, with ‘man/boy’ serving as the reference group. Sexual orientation (0 = straight/heterosexual, 1 = LGBTQ+, 2 = Asexual/Unsure) was dummy coded using SPSS with dummy variables created for all values except the first category, with heterosexual/straight serving as the reference group. The regression analyses were intended to describe patterns within this diverse cohort and were not designed for causal inference.

## Results

### Cohort description

Table 2 shows demographics of the sample. A total of 74 individuals between the ages of 10 and 24 completed the baseline questionnaire, with a mean age of 17.39 years *(SD* = 3.64). Of those that reported gender (*n* = 71), 33 (46.5%) identified as a woman/girl, 20 (28.2%) identified as a man/boy, 10 (14.1%) individuals identified as non-binary, and 8 (11.3%) participants identified as ‘other’ genders (e.g., agender, autigender, gender fluid). Nineteen participants (25.7%) identified as trans. In terms of ethnicity, the majority *(n =* 44, 59.5%) of individuals identified as White English, Welsh, Scottish, Northern Irish or British, followed by African (*n* = 6, 8.1%) and mixed or of multiple Ethnic background (*n* = 6, 8.1%), respectively. Most individuals *(n =* 60, 81.1%) were born in the UK and were in school or sixth form college (*n* = 36, 48.6%). Eight (10.9%) participants reported living in foster care (ever), and 14 (18.9%) reported experiencing hunger sometimes, once or twice. Over thirty percent (31.1%, *n* = 23) reported a formal neurodivergent diagnosis made by a clinician. The most common reported neurodivergent experiences (e.g., diagnosed and suspected but undiagnosed) included autism (*n* = 28,37.8%) followed by ADHD (*n* = 24, 32.4%).

**Table 2:**
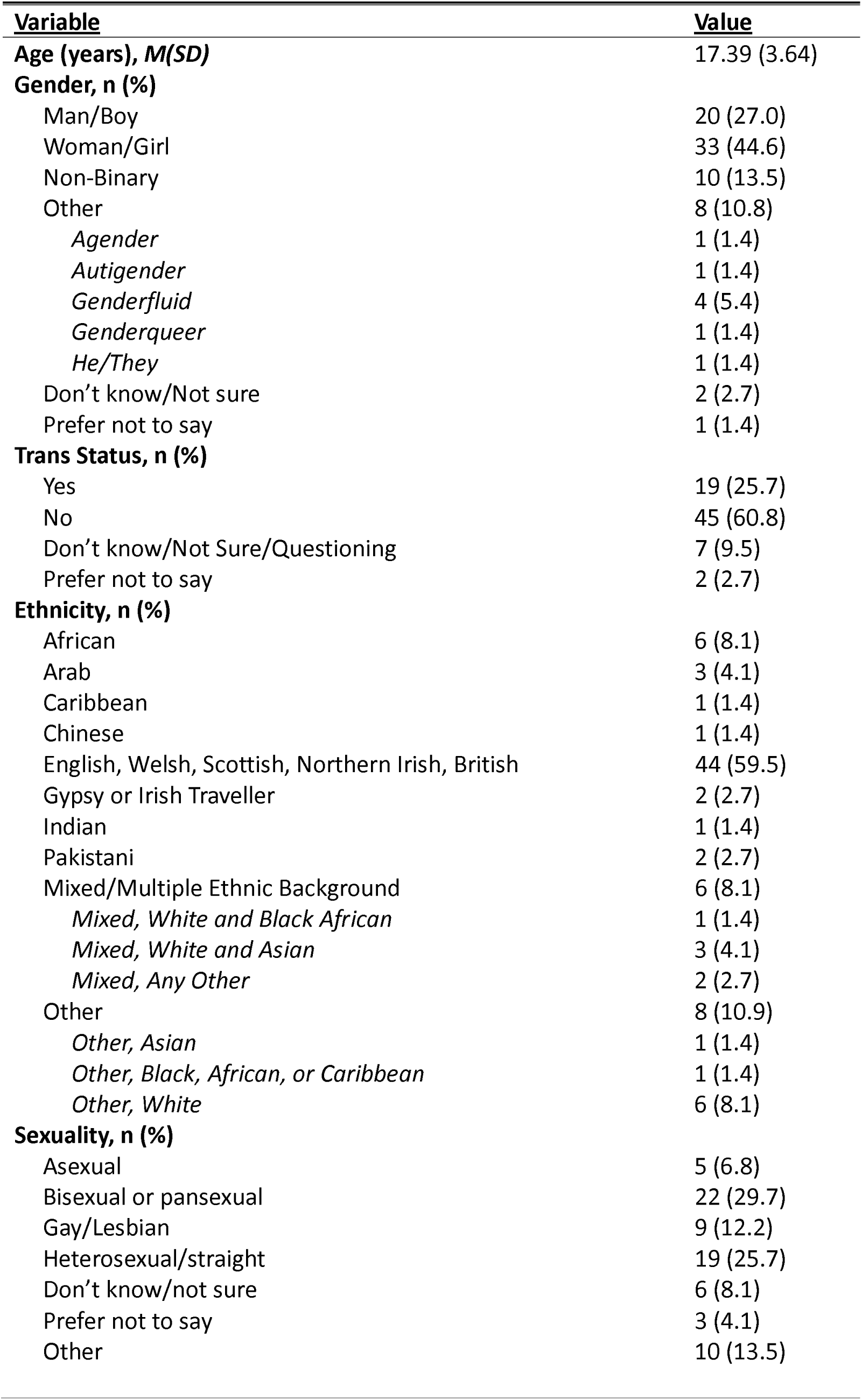
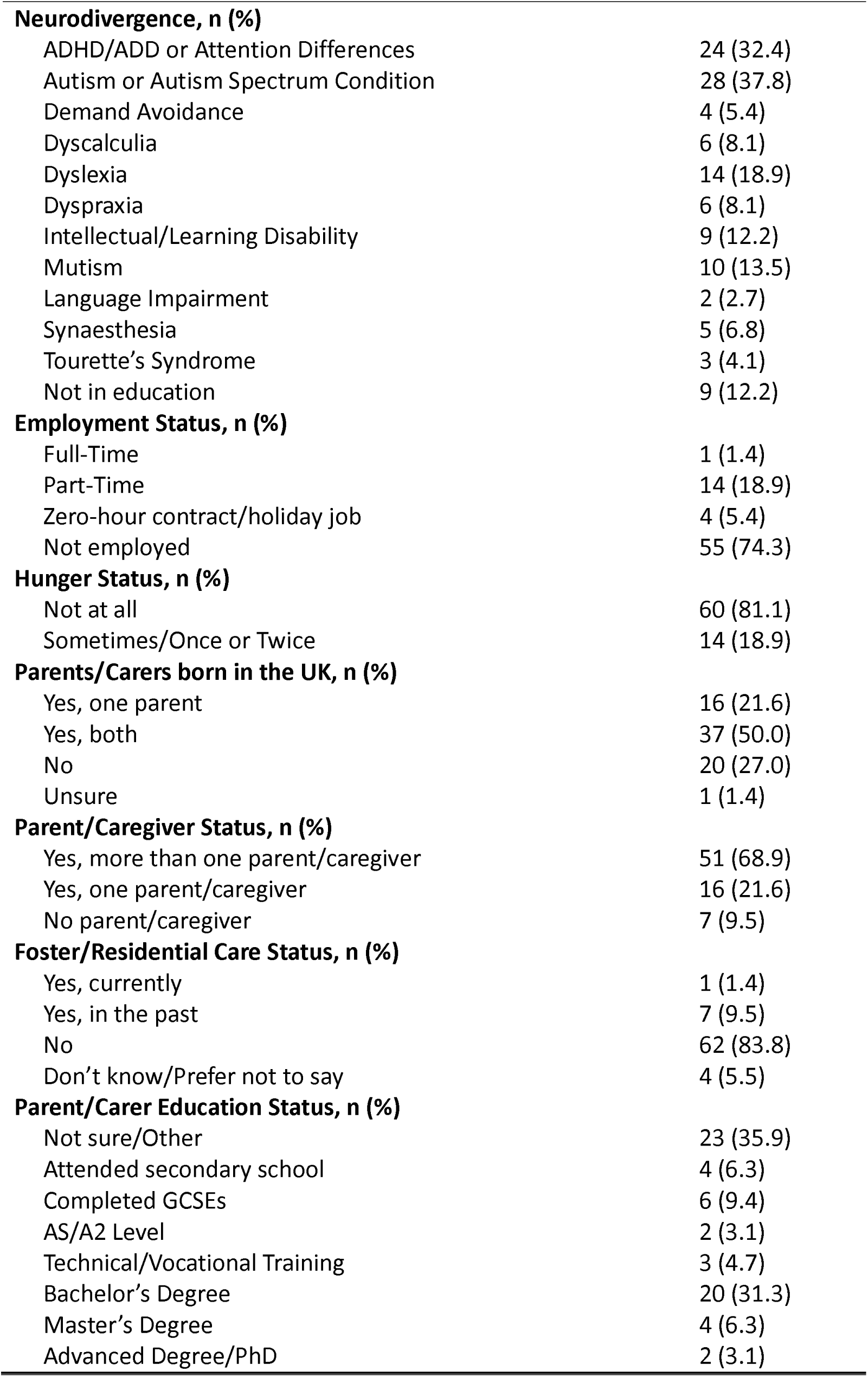
Sociodemographic Characteristics of Participants (*n* = 74).

| **Variable** | **Value** |
| --- | --- |
| **Age (years), M(SD)** | 17.39 (3.64) |
| **Gender, n (%)** |  |
| Man/Boy | 20 (27.0) |
| Woman/Girl | 33 (44.6) |
| Non-Binary | 10 (13.5) |
| Other | 8 (10.8) |
| *Agender* | 1 (1.4) |
| *Autigender* | 1 (1.4) |
| *Genderfluid* | 4 (5.4) |
| *Genderqueer* | 1 (1.4) |
| *He/They* | 1 (1.4) |
| Don't know/Not sure | 2 (2.7) |
| Prefer not to say | 1 (1.4) |
| **Trans Status, n (%)** |  |
| Yes | 19 (25.7) |
| No | 45 (60.8) |
| Don't know/Not Sure/Questioning | 7 (9.5) |
| Prefer not to say | 2 (2.7) |
| **Ethnicity, n (%)** |  |
| African | 6 (8.1) |
| Arab | 3 (4.1) |
| Caribbean | 1 (1.4) |
| Chinese | 1 (1.4) |
| English, Welsh, Scottish, Northern Irish, British | 44 (59.5) |
| Gypsy or Irish Traveller | 2 (2.7) |
| Indian | 1 (1.4) |
| Pakistani | 2 (2.7) |
| Mixed/Multiple Ethnic Background | 6 (8.1) |
| *Mixed, White and Black African* | 1 (1.4) |
| *Mixed, White and Asian* | 3 (4.1) |
| *Mixed, Any Other* | 2 (2.7) |
| Other | 8 (10.9) |
| *Other, Asian* | 1 (1.4) |
| *Other, Black, African, or Caribbean* | 1 (1.4) |
| *Other, White* | 6 (8.1) |
| **Sexuality, n (%)** |  |
| Asexual | 5 (6.8) |
| Bisexual or pansexual | 22 (29.7) |
| Gay/Lesbian | 9 (12.2) |
| Heterosexual/straight | 19 (25.7) |
| Don't know/not sure | 6 (8.1) |
| Prefer not to say | 3 (4.1) |
| Other | 10 (13.5) |
| **Neurodivergence, n (%)** |  |
| ADHD/ADD or Attention Differences | 24 (32.4) |
| Autism or Autism Spectrum Condition | 28 (37.8) |
| Demand Avoidance | 4 (5.4) |
| Dyscalculia | 6 (8.1) |
| Dyslexia | 14 (18.9) |
| Dyspraxia | 6 (8.1) |
| Intellectual/Learning Disability | 9 (12.2) |
| Mutism | 10 (13.5) |
| Language Impairment | 2 (2.7) |
| Synaesthesia | 5 (6.8) |
| Tourette's Syndrome | 3 (4.1) |
| Not in education | 9 (12.2) |
| **Employment Status, n (%)** |  |
| Full-Time | 1 (1.4) |
| Part-Time | 14 (18.9) |
| Zero-hour contract/holiday job | 4 (5.4) |
| Not employed | 55 (74.3) |
| **Hunger Status, n (%)** |  |
| Not at all | 60 (81.1) |
| Sometimes/Once or Twice | 14 (18.9) |
| **Parents/Carers born in the UK, n (%)** |  |
| Yes, one parent | 16 (21.6) |
| Yes, both | 37 (50.0) |
| No | 20 (27.0) |
| Unsure | 1 (1.4) |
| **Parent/Caregiver Status, n (%)** |  |
| Yes, more than one parent/caregiver | 51 (68.9) |
| Yes, one parent/caregiver | 16 (21.6) |
| No parent/caregiver | 7 (9.5) |
| **Foster/Residential Care Status, n (%)** |  |
| Yes, currently | 1 (1.4) |
| Yes, in the past | 7 (9.5) |
| No | 62 (83.8) |
| Don't know/Prefer not to say | 4 (5.5) |
| **Parent/Carer Education Status, n (%)** |  |
| Not sure/Other | 23 (35.9) |
| Attended secondary school | 4 (6.3) |
| Completed GCSEs | 6 (9.4) |
| AS/A2 Level | 2 (3.1) |
| Technical/Vocational Training | 3 (4.7) |
| Bachelor's Degree | 20 (31.3) |
| Master's Degree | 4 (6.3) |
| Advanced Degree/PhD | 2 (3.1) |

Descriptive data of mental health indicators and ACEs are presented in Table 3. More than a third of the sample (*n* = 26 participants, 36.1%) reported two or more maltreatment experiences, and 10% (n=8) reported four or more. The most common reported maltreatment was emotional abuse (*n* = 35, 47.5%), followed by emotional neglect *(n =* 25, 33.8%) and physical abuse *(n =* 20, 27.0%), respectively.

**Table 3:**
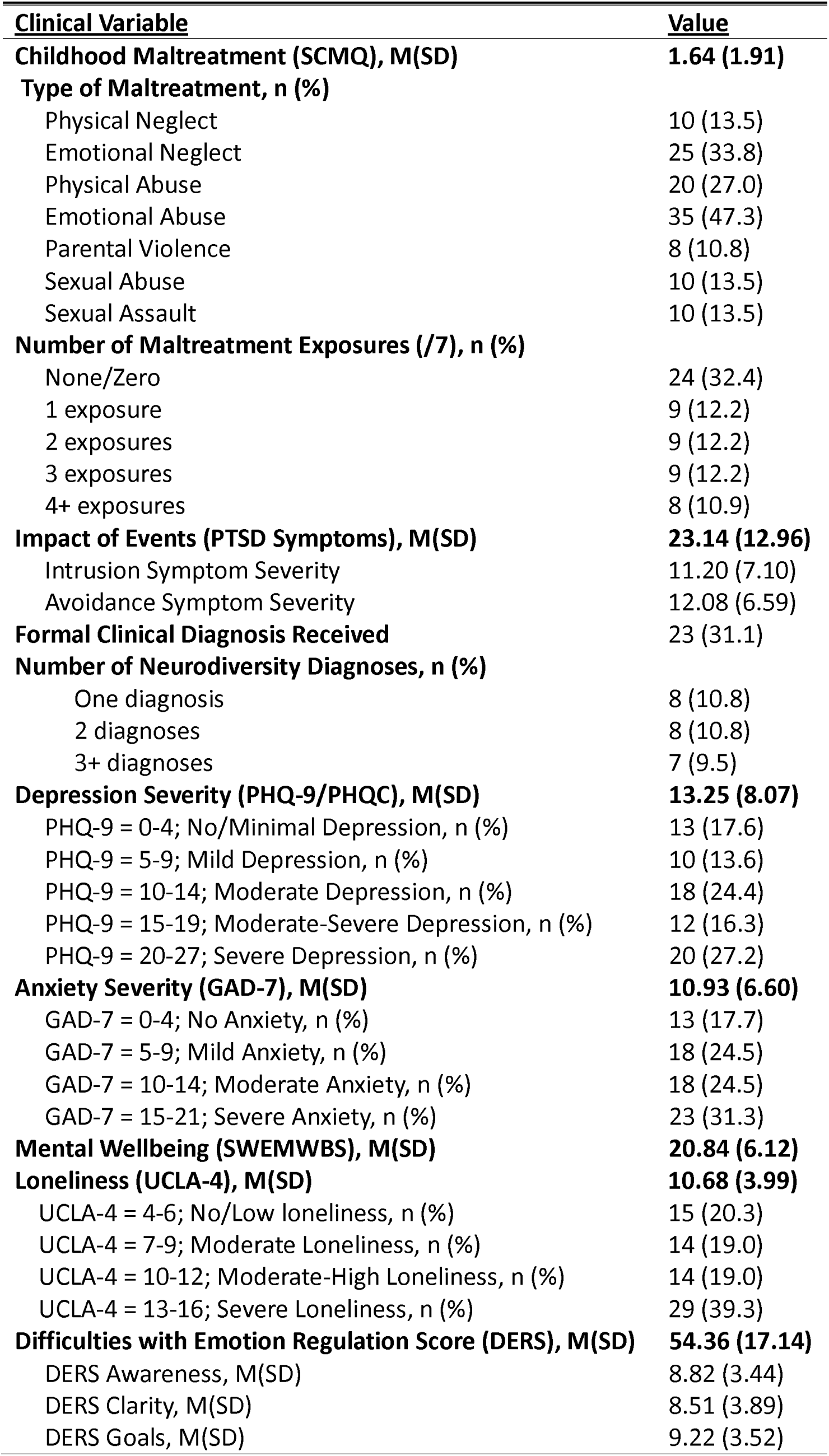
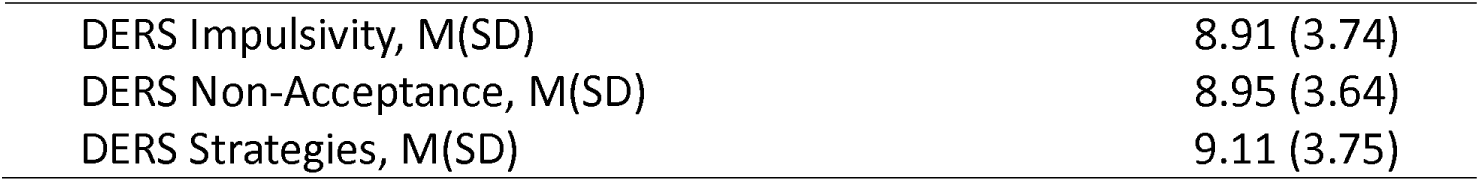
Childhood Maltreatment and Mental Health Characteristics of Participants (*n* = 74)

Mean depression (PHQ-9 *M =* 13.24, *SD =* 8.07) and anxiety (GAD-7 *M =* 10.93, *SD =* 6.60) scores were in the moderate severity range (PHQ9 and GAD scores of 10-14). Twenty participants (27.2%) reported severe depression (PHQ score of >20), and 23 participants (31.3%) reported severe anxiety (GAD7 score of >15). Of those who reported PTSD symptoms (*n* = 50), if the mean Children’s Revised Impact of Event Scale surpassed 17 (CRIES *M =* 23.14, *SD =* 12.96), this indicated a likely PTSD diagnosis.^18^

### Associations between ACEs, Mental Health Indicators, Social Position, Wellbeing, and Identity

Bivariate correlation analyses were conducted to examine patterns of association between number of ACEs, psychometric measures, and selected demographic variables (Table 4). Significant associations were observed between SCMQ scores (ACEs) and all mental health measures. The strongest associations were found for depression (PHQ-9 *r_p_ =* 0.53, *p* < .001), loneliness (UCLA *r_p_=* 0.56, *p <* .001) and wellbeing (SWEMWBS *r_p_* = −0.50, *p <* .001), in the expected directions. Higher ACEs exposure was associated with greater depression symptom severity, loneliness, and lower mental wellbeing. Clinical diagnosis of a neurodivergent experience and self-reported neurodivergent experiences were also correlated with depressive symptoms (PHQ-9 *r_p_* = .46, *p <* .001), anxiety symptoms (GAD-7 *r_p_* = .44, *p <* .001), difficulties with emotion regulation (DERS *r_p_ =* .44, *p <* .001), and inversely associated with wellbeing (SWEMWBS *r_p_ =* -.27, *p <* .05), again in the expected direction. (See additional preparatory analyses on mental health, ethnicity, gender identity, sexual orientation, trans identity in Supplementary File 1).

**Table 4:** Bivariate Correlation Coefficient Matrix of Associations between Childhood Maltreatment and Mental Health Outcomes (*N* = 74)

| Variable | *Variable/Correlation* |  |  |  |  |  |  |  |  |  |  |
| --- | --- | --- | --- | --- | --- | --- | --- | --- | --- | --- | --- |
|  | SCMQ | PHQ-9 | GAD-7 | CRIES | DERS | UCLA | SWEMWBS | NDxN | AGE | TRANS | ETH |
| 1. SCMQ | 1.00 |  |  |  |  |  |  |  |  |  |  |
| 2. PHQ-9 | .53** | 1.00 |  |  |  |  |  |  |  |  |  |
| 3. GAD-7 | .46** | .88** | 1.00 |  |  |  |  |  |  |  |  |
| 4. CRIES | .40** | .56** | .59** | 1.00 |  |  |  |  |  |  |  |
| 5. DERS | .49** | .82** | .75** | .56** | 1.00 |  |  |  |  |  |  |
| 6. UCLA | .56** | .59** | .54** | .45** | .53** | 1.00 |  |  |  |  |  |
| 7. SWEMWBS | -.50** | -.71** | -.59** | -.54** | -.77** | -.61** | 1.00 |  |  |  |  |
| 8. NDxN | .13 | .46** | .44** | .04 | .44** | .22 | -.27* | 1.00 |  |  |  |
| 9. AGE | .26* | .20 | .17 | .27 | .23 | .31** | -.23 | .24* | 1.00 |  |  |
| 10. TRANS | .19 | .39** | .41** | .08 | .29* | .11 | -.24* | .34** | -.09 | 1.00 |  |
| 11. ETH | -.03 | -.20 | -.27* | -.12 | -.15 | -.07 | .10 | -.19 | .03 | -.24* | 1.00 |
| 12. GEND | .04 | .27* | .36** | .10 | .10 | .22 | -.14 | .09 | .04 | .17 | .30* |
| 13. SEXOR | -.25 | -.45** | -.53** | -.40** | -.35** | -.29* | .30** | -.32** | .03 | -.53** | .44** |

**Note.** CRIES = Child Revised Impact of Events Scale; DERS = Difficulties in Emotion Regulation Scale; ETH = Ethnicity; GAD-7 = Generalized Anxiety Disorder Assessment; GEND = Gender; NDxN = Number of Neurodiversity Diagnoses and Experiences; PHQ-9 = Patient Health Questionnaire; SEXOR = Sexual Orientation; SCMQ = Short Child Maltreatment Questionnaire; SWEMWBS = Short Warwick-Edinburgh Mental Wellbeing Scale; TRANS = trans identity; UCLA = UCLA Loneliness Scale.

To examine associations between depressive symptom severity (PHQ-9), ACEs exposure, and identity characteristics, hierarchical multiple linear regression analyses were performed with PHQ-9 scores as the primary outcome (Table 5). In Model 1, sCMQ scores (ACEs) accounted for approximately 28% of the variance in PHQ-9 scores (p *< .001).* In Model 2, identity characteristics were entered simultaneously and explained a further 22% of the variance in PHQ-9 score. In the final three-predictor model, SCMQ scores (*B* = 1.86, *p <* .001), greater neurodivergent experiences (*B* = 1.44, *p =* .007), and non-binary gender (*B* = 4.86, *p =* .04) predicted higher baseline PHQ-9 scores *(F* = 7.12, *p <* .001, Adj. *R*^2^ = 0.43) accounting for 43% of the variance in PHQ-9 scores.

**Table 5:**
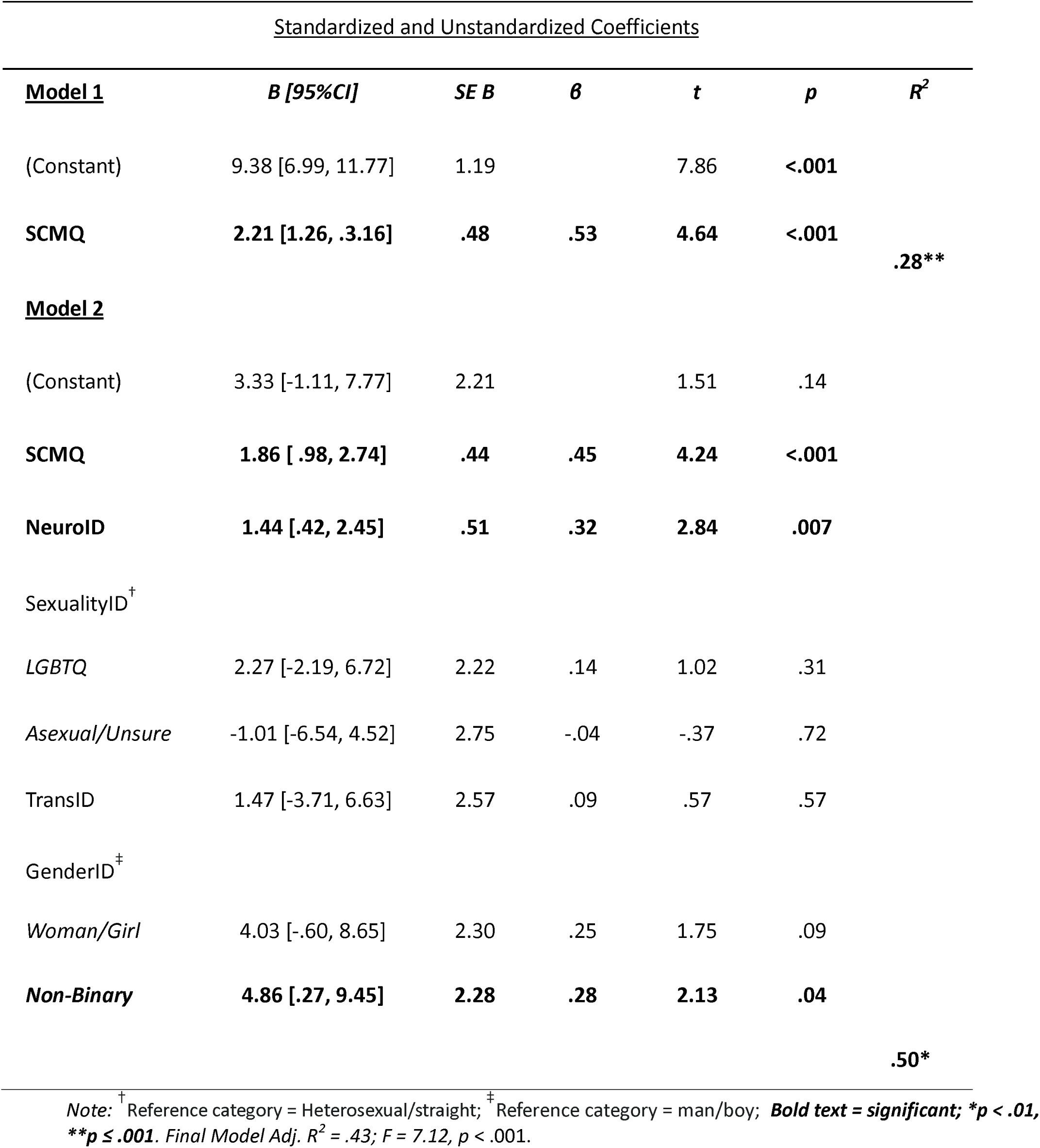
ummary of Multiple Linear Regression Analysis of Predictors of PHQ-9 Depression Score.

A hierarchical multiple linear regression analysis examined associations between ACEs exposure and selected identity variables with anxiety symptoms (GAD-7; see Table 6). In Model 1, SCMQ (ACEs) score was significantly associated with higher GAD-7 anxiety scores, explaining 23% of the variance (*B* = 1.55, *p < .001).* In Model 2, the addition of identity characteristics resulted in a further 27% increase in explained variance in GAD-7. In the final model, higher SCMQ scores (*B* = 1.28, *p <* .001), greater number of neurodivergent experiences (*B* = .87, *p =* .04), and non-binary gender identity (*B* = 4.75, *p* < .01), significantly predicted higher GAD-7 scores *(F* = 5.95, *p <* .001, Adj. *R*^2^ = 0.42) and accounted for 42% of the variance in anxiety symptoms.

**Table 6:** Summary of Multiple Linear Regression Analysis of Predictors of GAD-7 Anxiety Score.

| <u>Standardized and Unstandardized Coefficients</u> |
| --- |

| **Model 1** | ***B [95%CI]*** | ***SE B*** | ***β*** | ***t*** | ***p*** | ***R*<sup>2</sup>** |
| --- | --- | --- | --- | --- | --- | --- |
| (Constant) | 8.28 [6.29, 10.16] | .99 |  | 8.36 | <.001 |  |
| **SCMQ** | **1.55 [.76, 2.33]** | **.39** | **.47** | **3.96** | **<.001** | **.23\*\*** |
| **Model 2** |  |  |  |  |  |  |
| (Constant) | 5.28 [-1.03, 11.59] | 3.14 |  | 1.68 | .10 |  |
| **SCMQ** | **1.28 [.58, 1.99]** | **.35** | **.39** | **3.65** | **<.001** |  |
| **NeuroID** | **.87 [.05, 1.69]** | **.41** | **.25** | **2.15** | **.04** |  |
| SexualityID<sup>†</sup> |  |  |  |  |  |  |
| *LGBTQ+* | 2.20 [-1.78, 6.18] | 1.98 | .17 | 1.11 | .27 |  |
| *Asexual/Unsure* | -1.04 [-5.56, 3.78] | 2.40 | -.06 | -.43 | .67 |  |
| TransID | 1.20 [-3.06, 5.45] | 2.12 | .09 | .57 | .58 |  |
| GenderID<sup>‡</sup> |  |  |  |  |  |  |
| *Woman/girl* | 3.31 [-.59, 7.21] | 1.94 | .26 | 1.71 | .09 |  |
| ***Non-Binary*** | **4.75 [1.07, 8.43]** | **1.83** | **.45** | **2.60** | **.01** |  |
| EthnicID | -1.58 [-4.96, 1.79] | 1.68 | -.12 | -.94 | .35 |  |
|  |  |  |  |  |  | **.50\*\*** |

*Note:* <sup>†</sup> Reference category = Heterosexual/straight; <sup>‡</sup> Reference category = man/boy; **Bold text = significant; \**p* < .05, \*\**p* ≤ .01. Final Model Adj. *R*<sup>2</sup> = .42; *F* = 5.95, *p* < .001.**

## Discussion

Using inclusive, co-designed, and trauma-informed recruitment methods, we demonstrated the feasibility of collecting mental health and adversity data from groups often excluded from research due to concerns about distress, engagement, and perceived vulnerability. The observed cross-sectional associations between adverse childhood experiences and mental health outcomes are consistent with prior research demonstrating the enduring impact of childhood adversity on mental and emotional health. While causal inferences cannot be drawn, these findings suggest that established patterns are also evident within cohorts of young people who have typically been underrepresented in ACE research.^12^

Higher exposures to ACES, a greater number of neurodivergent experiences and identifying as non-binary gender independently associated with greater depression and anxiety symptom severity. The association between the number of neurodivergent experiences and poorer mental health suggests that neurodivergent young people face unique psychological and social challenges that are important to consider. This may reflect the interaction between neurodivergent ways of processing and environments not designed to accommodate them, alongside broader systemic challenges related to stigma, exclusion, unmet support needs, and misunderstanding. Importantly, our findings point to the value of inclusive research practices that ensure neurodivergent young people are represented and their experiences centred in mental health research.

Identifying as non-binary was associated with higher depression and anxiety symptom severity compared with men/boys, possibly due to increased exposure to discrimination, stigma and lack of affirming social support, all of which are linked with poorer mental health outcomes.^23^ ^24^ Our findings highlight that including expanded assessments of gender identities beyond the typical binary category (i.e., man/boy vs. woman/girl) explains significant additional variance in mental health outcomes.

No significant differences between men/boys and women/girls were found, contradicting previous research suggesting that women/girls are up to twice as likely to have mental health challenges.^25^ These gender differences are known to emerge after puberty and later adolescence, so the wide age range of the present cohort, combined with multiple sources of disadvantage and adversity may have influenced our findings.

### Study Limitations

The additional support and co-codesign process took much time, perhaps making large future cohort studies and trials of such diverse young people harder to resource. The relational and procedural supports required for meaningful inclusion may constrain rapid scale-up but enhance ethical integrity and epistemic validity. This argues for more participatory, creative, and co-designed recruitment approaches for ultra-marginalised groups to ensure that their experiences inform policy and practice.

The sample size of 74 adolescents limits statistical power, particularly for subgroup analyses, and for estimating intersectional effects and interactions (not conducted in this study). Importantly, sexual orientation, transgender identity specifically, and ethnicity did not emerge as significant correlates of depression or anxiety. This could potentially be explained by methodological limitations (e.g., small sample of those who identified as trans or LGBTQ+ and aggregation of the variables for meaningful statistical analyses may have obscured sub-group specific effects).

Furthermore, *protective sampling* by community partners might have led to those with the greatest needs not entering the cohort. The study was conducted within a single country, which may limit the generalisability of the findings to adolescents in other cultural, social, or economic contexts, especially those from low and middle-income countries.

### Conclusions

This study demonstrates inclusive research methods to recruit and engage a highly marginalised cohort of young people across England. Our findings revealed coherent patterns of associations between ACEs, neurodivergence, gender identity, and mental health outcomes. Future studies should test these methods to recruit larger samples and longitudinal quantitative and qualitative and creative co-designs.

## Ethical Information

Informed consent and assent of participants was obtained from all participants prior to completion of any assessments. The study was reviewed and approved in 2022 by the University of Oxford’s Research Ethics Committee (R71941/RE001) and the Health Research Authority (23/WM/0105).

## Author Contributions

KB, M.K. contributed equally as co-first authors. KB acquired the funding: conceptualisation, supervision, ethics, administration, and programme delivery, reconciling contributions and producing a final version with MK. I.B. Conceptualisation, investigation, methodology, project administration, writing -original draft, writing- review and editing. Data curation, analysis, investigation, methodology. M.F.: Writing - review and editing. P.C., C.F., A.F., K.H., H.S., N.S., S.H-J., L.A., A.M. LH.: statistical advice, writing - review and editing.

## Conflicts of Interest Declared

KB is Deputy Editor for BJPsych Open and former Editor in Chief BJPsych. KB has played no role in the processing or decisions on this paper. There are no other conflicts of interest to declare.

## Data Availability Statement

The data that supports the findings of this study are available in the supplementary material of this article.

## Funding Received

This work was supported by the UKRI Medical Research Council. (MR/ W002183/1). MKC and KB are funded by the National Institute for Health and Care Research (NIHR), Oxford Health Biomedical Research Centres (BRC). The funders had no role in designing the study, the analysis, or the decision to submit the paper. The views expressed are those of the authors and not necessarily those of the NIHR or the Department of Health and Social Care.

## Acknowledgements

The authors wish to thank each member of the wider ATTUNE research team who has helped in the design, conceptualisation of the project including those delivering those delivering the workshops in each location and the broader team.

