## Supplementary File for "Assessing adverse childhood experiences and mental health status in diverse and underrepresented young people: advancing Inclusive research"

*Table 1. All Measures included in the ATTUNE Programme survey*

| General demographics and characterisation | Region, age, gender, sexual orientation, ethnicity, religion, care experiences, education and employment status |
| --- | --- |
| Neurodivergence | Self-identified and diagnosed differences in experience, including ADHD, autistic spectrum condition and learning difficulties (ATTUNE-designed) |
| ACEs | Short Child Maltreatment Questionnaire |
| Mental health and wellbeing | Generalised Anxiety Disorder Assessment (GAD -7)  Patient Health Questionnaire (PHQ-9 or Child-PHQ-9) Warwick Edinburgh Mental Wellbeing Scale (WEMWBS -7) UCLA Loneliness scale, 4-item Russell 1996 version (ULS-4) |
| Emotion regulation | Difficulties in Emotion Regulation Scale – Short Form (DERS)  Revised Impact of Events Scale -13 (CRIES-13 or Child CRIES-13) |

**Exploratory Analysis of Sociodemographics and Mental Health Outcomes**

To explore the associations between ethnicity and mental health outcomes, an independent *t-*test was performed to detect differences in psychometric outcomes based on ethnicity (Table 2). Findings revealed that participants who identified as White had significantly higher anxiety scores (12.08 ± 6.54) compared to those that identified as non-White (8.14 ± 5.99), *t*(70) = *2.75, p* = .02. No other significant differences in social and mental health outcomes based on ethnicity were found.

**Table 2.** *Associations between ethnicity and mental health indicators (n = 74)*

| Variable,*M (SD)* | **White (*n* = 52)** | **Non-White (*n* = 22)** | *p* |
| --- | --- | --- | --- |
| **PHQ-9** | 14.25 (7.91) | 10.76 (8.12) | .09 |
| **GAD-7** | 12.08 (6.54) | 8.14 (5.99) | **.02*** |
| **SWEMWBS** | 20.45 (5.61) | 21.73 (7.22) | .42 |
| **UCLA** | 10.86 (4.14) | 10.27 (3.68) | .57 |
| **SCMQ** | 1.68 (1.85) | 1.56 (2.09) | .82 |
| **CRIES** | 24.18 (13.7) | 20.94 (11.32) | .42 |
| **DERS** | 55.94 (16.85) | 50.06 (17.68) | .22 |

Table 3 summarises the one-way ANOVAs conducted on gender identity and mental health indicators. The findings revealed a statistically significant difference based on gender identity in PHQ-9 depression scores (*F(2,72) =* 4.57*, p* = .01) and GAD-7 scores (*F*(2, 71) = 7.91, *p* <.001). A Tukey HSD post hoc comparison revealed that non-binary/other identity was associated with significantly higher GAD-7 anxiety scores (*M =* 15.33, *SD =* 4.20) compared to those that identified as a man/boy (*M =* 9.32, *SD =* 6.98, *p* = .01) and woman/girl (*M =* 9.00, *SD =* 6.43, *p* = .001). PHQ-9 depression scores were significantly higher for non-binary/other participants (*M =* 17.52, *SD =* 6.54) compared to man/boy (*M =* 11.75, *SD* = 8.37, *p* = .05) and woman/girl (*M =* 11.38, *SD* = 7.95, *p* = .02). No other significant differences based on gender identity were found (*p* > .05).

**Table 3.** *Association between gender identity and mental health indicators (n = 73)*

| Variable  *M (SD)* | **Man/Boy (*n* = 20)** | **Woman/Girl (*n* = 32)** | **Non-Binary/ Other**  **(*n* = 21)** | ***F*** | ***p*** |
| --- | --- | --- | --- | --- | --- |
| **PHQ-9** | 11.75 (8.37) | 11.38 (7.95) | 17.52 (6.54) | **4.57** | **.01** |
| **GAD-7** | 9.32 (6.98) | 9.00 (6.43) | 15.33 (4.20) | **7.91** | **<.001** |
| **SWEMWBS** | 21.42 (6.91) | 21.78 (6.33) | 18.81 (4.63) | 1.67 | .20 |
| **UCLA** | 10.26 (4.00) | 9.81 (3.91) | 12.38 (3.72) | 2.93 | .06 |
| **SCMQ** | 1.88 (2.26) | 1.44 (1.96) | 1.71 (1.49) | .28 | .76 |
| **CRIES** | 20.62 (14.03) | 23.79 (13.01) | 24.46 (12.43) | .34 | .72 |
| **DERS** | 54.17 (18.08) | 51.11 (18.06) | 58.86 (14.67) | 1.24 | .30 |

Independent *t-*tests were performed to detect differences in psychometric outcomes based on trans identity. Findings revealed that participants who identified as trans or were questioning/unsure (*n =* 26) had significantly higher PHQ-9 depression scores (*t(*70) = -3.51, *p* < .001, *d* = -.86), GAD-7 anxiety scores (*t(*69) = 3.72, *p* < .001, *d* = -.92), SWEMWBS wellbeing scores (*t(*70) = 2.08, *p* = .02, *d* = .51), and DERS scores (*t(*64) = -2.46, *p* .008, *d* = -.62), compared to those that identified as not trans. No other significant differences in social and mental health outcomes based on trans identity were found (Table 4).

**Table 4:** Associations between trans identity and mental health indicators (*n* = 72)

| Variable, *M (SD)* | **Trans/Questioning**  **(*n* = 26)** | **Not Trans/DNS**  **(*n* = 46)** | ***p*** | ***d*** |
| --- | --- | --- | --- | --- |
| **PHQ-9** | 17.38 (6.44) | 10.89 (8.10) | **<.001** | **-.86** |
| **GAD-7** | 14.54 (4.80) | 8.98 (6.69) | **<.001** | **-.92** |
| **SWEMWBS** | 18.88 (5.22) | 21.96 (6.43) | **.02** | **.51** |
| **UCLA** | 11.27 (3.94) | 10.38 (4.06) | .19 | -.22 |
| **SCMQ** | 2.10 (2.11) | 1.35 (1.77) | .08 | -.39 |
| **CRIES** | 24.65 (11.81) | 22.36 (13.62) | .28 | -.18 |
| **DERS** | 60.54 (15.43) | 50.25 (17.36) | **.008** | **-.62** |

**Table 5** summarises the one-way ANOVAs exploring differences in mental health indicators based on sexual identity. The results revealed statistically significant differences based on sexual orientation in PHQ-9 depression scores (*F(3,69) = 6.51, p* < .001), GAD-7 anxiety scores (*F*(3, 68) = 9.33, *p* <.001), CRIES PTSD score (*F(3,46) =* 3.87*, p* = .02), SWEMWBS wellbeing scores (*F(3,69) =* 4.42*, p* = .007), UCLA Loneliness Scale (*F(3,89) =* 2.73*, p* = .05), and DERS emotion dysregulation scores (*F(3,63) =* 4.25*, p* = .008). Tukey HSD post-hoc comparisons revealed that LGBTQ+ was associated with significantly higher depression, anxiety, worsening wellbeing, and greater dysregulation (*p* < .05).

**Table 5:** Sexual orientation and Social/Mental Health Outcomes (*n* = 63)

| Variable  *M (SD)* | **LGBTQ+ (*n* = 41)** | **Asexual/Unsure**  **(*n* = 11)** | **Straight (*n* = 18)** | **Prefer not to Say**  **(n = 3)** | ***F*** | ***p*** |
| --- | --- | --- | --- | --- | --- | --- |
| **PHQ-9** | 16.56 (6.41) | 9.55 (9.82) | 8.83 (7.47) | 8.00 (7.00) | **6.51** | **<.001** |
| **GAD-7** | 13.97 (5.18) | 7.36 (7.89) | 7.00 (5.34) | 4.67 (4.16) | **9.33** | **<.001** |
| **SWEMWBS** | 18.85 (4.89) | 25.40 (6.41) | 22.68 (6.59) | 21.00 (8.72) | **4.42** | **.007** |
| **UCLA** | 11.66 (3.75) | 8.80 (4.52) | 10.11 (3.89) | 7.00 (1.00) | **2.73** | **.05** |
| **SCMQ** | 2.09 (2.08) | 1.00 (1.60) | 1.18 (1.63) | 1.00 (1.41) | 1.33 | .27 |
| **CRIES** | 27.67 (11.25) | 17.28 (12.87) | 20.07 (13.18) | 4.00 (5.66) | **3.87** | **.02** |
| **DERS** | 59.56 (15.12) | 42.78 (17.20) | 49.33 (17.79) | 37.50 (13.44) | **4.25** | **.008** |
